# Confidence in perceptual decision-making is preserved in schizophrenia

**DOI:** 10.1101/2019.12.15.19014969

**Authors:** Nathan Faivre, Matthieu Roger, Michael Pereira, Vincent de Gardelle, Jean-Christophe Vergnaud, Christine Passerieux, Paul Roux

**Affiliations:** Université Grenoble Alpes, CNRS, LPNC UMR 5105, Grenoble, France; Centre d’Economie de la Sorbonne, CNRS UMR 8174, Paris, France; Université Versailles Saint-Quentin-En-Yvelines, EA 4047 HandiResp, Montigny-Le-Bretonneux, France; Centre Hospitalier de Versailles, Service Hospitalo-Universitaire de psychiatrie d’adultes et d’addictologie, Le Chesnay, France

**Keywords:** confidence, schizophrenia, metacognition, cognitive insight, depression, trajectory tracking, evidence accumulation model

## Abstract

Metacognition is the set of reflexive processes allowing humans to evaluate the accuracy of their mental operations. Deficits in synthetic metacognition have been described in schizophrenia using mostly narrative assessment and linked to several key symptoms. Here, we assessed metacognitive performance by asking individuals with schizophrenia or schizoaffective disorder (N=20) and matched healthy participants (N = 21) to perform a visual discrimination task and subsequently report confidence in their performance. Metacognitive performance was defined as the adequacy between visual discrimination performance and confidence. Bayesian analyses revealed equivalent metacognitive performance in the two groups despite a weaker association between confidence and trajectory tracking during task execution among patients. These results were reproduced using a bounded evidence accumulation model which showed similar decisional processes in the two groups. The inability to accurately attune confidence to perceptual decisions in schizophrenia remains to be experimentally demonstrated, along with the way such impairments may underpin functional deficits.

## Introduction

Metacognition refers to a spectrum of mental activities whose objects are the subject’s own thoughts. Some of these mental activities can be described as discrete (recognition and monitoring of ongoing thoughts or percepts), others as more transversal and synthetic, integrating a subject’s assumption of thoughts, sensations, intentions or links between events to form more complete and lasting representations (Lysaker et al., 2013 ; Bob et al., 2016 ; David, 2019). Regarding the latter, individuals with schizophrenia have persistent difficulties in considering thoughts as essentially subjective, in recognizing complex mental states in others, in viewing events from perspectives other than their own, and in using their metacognitive knowledge to manage their distress (Hasson-Ohayon et al., 2015; Lysaker & Dimaggio, 2014). These deficits have been linked to core features of the illness like positive and negative symptoms (McLeod, Gumley, MacBeth, Schwannauer, & Lysaker, 2014), disorganisation (Vohs et al., 2014), functioning (Lysaker et al., 2010), and quality of life (Arnon-Ribenfeld, Hasson-Ohayon, Lavidor, Atzil-Slonim, & Lysaker, 2017). Synthetic metacognition is usually measured through structured interviews and self-reported questionnaires that evaluate multiple processes such as emotion recognition, theory of mind and verbal abilities. In contrast, discrete metacognition is measured by focusing on a specific cognitive domain: participants are asked to perform a cognitive or perceptual task (sometimes referred to as the first-order task), and subsequently assess how well they performed (i.e., a second-order task consisting of a confidence judgment, error detection, or post-decisional wagering). In this context, metacognitive performance is defined as the capacity to adapt second-order judgments to first-order performance (Fleming & Lau, 2014).

Studies relying on such combinations of first and second-order tasks reported deficits in metacognitive performance in schizophrenia across several domains, such as perception (Moritz, Woznica, Andreou, & Kother, 2012), agency (Metcalfe, Van Snellenberg, DeRosse, Balsam, & Malhotra, 2012) and memory (Moritz, Woodward, Jelinek, & Klinge, 2008). Although these studies have provided valuable insights regarding putative deficits in discrete metacognition, several biases might interfere with the assessment of metacognitive performance in schizophrenia. First, it is important to consider that metacognitive performance depends on first-order performance: it is easier to provide confidence judgments or detect errors for simple than for difficult tasks. Thus, it is crucial to control for first-order task performance, which is usually lower in schizophrenia compared to controls. Other biases might influence metacognition in schizophrenia such as depression which has been associated with better metacognition (Lysaker et al., 2005) and cognitive deficits that have been associated with metacognitive impairments with a small-to-moderate effect size (Davies & Greenwood, 2018). Considering the many stages of processing leading from first to second-order decisions, poor metacognitive performance in a given task may be due to deficits at any of these levels.

Here, we sought to pinpoint the putative origins of metacognitive deficits in schizophrenia and describe how first and second-order cognitive processes unfold over time by analyzing behavioral responses together with trajectory tracking, and by reproducing them using a bounded evidence accumulation model of decision-making. Namely, we continuously tracked the position and kinematics of the mouse that participants used to indicate their first-order response during a motion discrimination task (Dotan, Meyniel, & Dehaene, 2018; Dotan, Pinheiro-Chagas, Al Roumi, & Dehaene, 2019). In addition, we modeled first and second-order responses as derived from a bounded evidence accumulation process starting when participants initiated a mouse movement (Pereira et al., 2018; Pleskac, Busemeyer, & others, 2010; Resulaj, Kiani, Wolpert, & Shadlen, 2009; Van Den Berg et al., 2016). Together, these two approaches following a pre-registered plan allowed us to finely characterize decision-making and metacognitive monitoring in schizophrenia in relation to clinical traits while avoiding the typical confounds that may have contaminated previous results in the field.

## Results

### 1. First and second-order performance

Twenty-three healthy volunteers (15 males, 8 females) from the general population and twenty individuals with a schizophrenia spectrum disorder (16 males, 4 females) took part in this study. Two healthy volunteers were excluded from the analysis, respectively due to a convergence failure during the staircase procedure, and an estimated IQ < 70. On each trial, participants were asked to indicate the mean motion direction of a random-dot kinetogram (RDK) by clicking within a circular frame located on the top, to the right or to the left of the stimulus (first-order task), and report how confident they were in their response (second-order task, see methods). The analysis of first-order task performance revealed that individuals with schizophrenia had a tendency to judge the RDK as moving rightward more often (criterion schizophrenia: −0.32 ± 0.15, criterion controls: 0.00 ± 0.21, t(37.7) = 2.50, p = 0.02, BF = 3.24). However, the two groups had similar discrimination performance (d’ schizophrenia: 1.29 ± 0.08, d’ controls: 1.38 ± 0.11, t(36.7) = 0.85, p = 0.40, BF = 0.41). The variance in motion direction corresponding to such performance as titrated with an adaptive procedure (see methods) was higher in controls (2.01 ± 0.20) than in patients (1.59 ± 0.17, t(38.9) = 3.09, p = 0.004, BF = 10.60), indicating that patients had reduced perceptual abilities.

At the second-order level, average confidence ratings were similar between groups (schizophrenia: 0.71 ± 0.05; controls: 0.70 ± 0.06, t(38.4) = 0.12, p = 0.91, BF = 0.31), as well as confidence bias defined as B-ROC (patients: −1.93 ± 0.26; controls: −2.06 ± 0.21, t(36.5) = 0.54, p = 0.59, BF = 0.35). We then estimated metacognitive efficiency (i.e., ratio between d’ and meta-d’) to capture the amount of perceptual evidence used by participants when computing confidence estimates. We made the prior assumption that controls had higher metacognitive efficiency (i.e., prior with Gaussian distribution of mean = 0.2 and SD = 1), based on the difference in metacognitive accuracy between first-episode psychosis and healthy controls recently reported by Davies and colleagues (2018). Results showed that the two groups had similar metacognitive efficiency (schizophrenia: 0.52, highest posterior density interval = [0.40 0.65], controls: 0.49, highest posterior density interval = [0.37 0.64]), with a Bayes factor of 0.18 supporting the absence of difference between groups (Figure 2B). Another metric of metacognitive performance was computed, namely metacognitive sensitivity which corresponds to the slope of the logistic regression between first-order accuracy and confidence. A similar prior assumption for higher metacognitive efficiency in the control group was made, represented by a steeper slope (i.e., Gaussian distribution with mean = 1, SD = 5). We chose a weakly informative prior in the absence of published evidence. No interaction between group and confidence was found (estimate = 0.06, highest posterior density interval = [−0.15 0.27], Bayes factor = 0.02) (Figure 2C). Importantly, Bayes factors smaller than 0.3 both for metacognitive efficiency and sensitivity support the null hypothesis, according to which individuals with schizophrenia have no impairment while adjusting confidence to their performance.

Following our pre-registered plan, we then sought to assess how motor behavior related to first-order responses modulated confidence ratings. As a first step, we quantified the relationship between confidence, first-order accuracy and standardized reaction times between groups using a mixed-effects linear regression including perceptual evidence as a regressor of interest. We found a negative relationship between confidence and standardized reaction times (estimate = −0.05 [−0.07 −0.04], evidence ratio > 4000), which indicates that confidence was high following fast first-order responses. This relationship was modulated by first-order accuracy (interaction accuracy * reaction times: estimate = −0.01 [−0.02 −0.01], evidence ratio = 221.22) and group (interaction group * reaction times: estimate = 0.02 [0.00 0.04], evidence ratio = 22.26) indicating that the slope between confidence and standardized reaction times was steeper for correct responses and for the control group. Interestingly, a similar pattern was found for standardized second-order reaction times, with a main effect (estimate = −0.02 [−0.03 −0.01], evidence ratio = 71.73) which was stronger in the control vs. schizophrenia group (interaction group * second-order reaction times: estimate = 0.02 [0.00 0.04], evidence ratio = 12.51). Together, these results indicate that standardized first and second-order reaction times covary with confidence to a lesser extent in individuals with schizophrenia, suggesting they may rely less on this input to form confidence estimates. Of note, raw first and second-order reaction times per se did not differ between groups (t(39.0) = 1.07, p = 0.29, BF = 0.49 and t(36.0) = 1.41, p = 0.17, BF = 0.66, respectively).

### 2. Trajectory tracking

Beyond reaction times, we quantified how mouse trajectories leading to first-order responses predicted subsequent confidence judgments (see Figure 2A for raw trajectories). First, we isolated trials in which a change of mind occurred, that is when participants started moving towards one response circle and later changed direction towards the other (see methods). Changes of mind corresponded respectively to 7.9 ± 2.7 % and 7.3 ± 2.9 % of total trials in the patient and control groups (t(37.9) = 0.32, p = 0.75, BF = 0.32). Interestingly, a mixed-effects logistic regression revealed that changes of mind were associated with lower first-order accuracy in both groups (main effect: estimate = −0.32, z = −2.07, p = 0.04, see Figure 2B), without significant interaction between group and accuracy (estimate = 0.18, z = 0.84, p = 0.40). Conversely, a mixed-effects linear regression revealed that changes of mind were associated with lower confidence (F(1,30.3) = 32.05, p < 0.001), and that this decrease was more pronounced in controls vs. patients (interaction term: F(1,30.33) = 5.03, p = 0.03). Together, these findings suggest that patients revised less their confidence following changes of mind that lead to errors. Of note, the relationships between confidence, first-order accuracy and standardized reaction times were similar to the ones mentioned above when excluding trials with changes of mind.

Next, we assessed how the slopes of individual trajectories covaried with confidence. We found a negative relationship between slopes and confidence (F(1,50.5) = 5.7, p = 0.02), independent of groups and first-order accuracy (Figure 2A). This suggests that both patients and controls moved the mouse more laterally for responses associated with high confidence, whether correct or not. In addition, we fitted a linear model to individual trajectories and found a positive relationship between the goodness of fit (R^2^) and confidence (F(1,40.1) = 17.5, p < 0.001) independent of group and first-order accuracy, revealing that confidence ratings were higher after responses following more linear trajectories. Of note, a trend suggested lower R^2^ in the patient group (F(1,37.1) = 3.73, p = 0.06). Besides spatial trajectories, we quantified how velocity and acceleration profiles related to confidence, by fitting mixed-effects linear regressions for each time sample across individual trials, with confidence and group as fixed effects. Of note, we centered data to zero to account for potential motor impairment in schizophrenia (Manschreck, Maher, Rucklos, & Vereen, 1982). For velocity, we found a main effect of confidence indicating that velocity reached higher peaks in high confidence trials, and an interaction between confidence and groups indicating that the positive correlation between velocity and confidence was significant in the two groups, but stronger in the control than in the schizophrenia group (p < 0.05 fdr-corrected, Figure S1). This interaction was explored by fitting velocity models for each group, which showed a sustained correlation between confidence and velocity at movement onset and offset among the control group, and a short-lived correlation at movement onset in the patient group (Figure 2D). For acceleration, we found a main effect of confidence, by which acceleration at movement onset reached higher values in high confidence trials, and an interaction between confidence and group close to movement offset, by which movement acceleration reached more negative values for healthy controls in high confidence trials (p < 0.05 fdr-corrected figure S1). As for velocity, this interaction was explored by fitting acceleration models for each group, which showed that the correlation between confidence and acceleration was significant both at movement onset and offset among the control group (Figure 2D). Among patients, only a weaker correlation following movement offset was found. Together, these results confirm the existence of kinematics correlates of confidence at the motor execution stage, and suggest that they may be stronger predictors of confidence in healthy individuals compared to schizophrenia patients.

To compare the underlying decision mechanisms between groups, we fitted a bounded evidence accumulation model (Vickers, 1977; Usher & McClelland, 2001; Kiani & Shadlen, 2009; van den Berg et al., 2016) to movement onset timings and the accuracy of the initial direction of the mouse trajectories. The decision process was modeled using two anticorrelated race accumulators (one accumulating evidence for left choices and one for right choices). Confidence ratings were simulated by extending the evidence accumulation process after the initial decision (Pleskac et al., 2010). The model comprised five free parameters. The *non-decision time* was the sum of the time needed before visual information started being integrated by the evidence accumulation process and the delay between the decision and the actual movement initiation. The *decision bound* corresponded to the amount of evidence needed to initiate a movement and the *drift rate* to the average slope of the evidence accumulation process. Finally, two *confidence criteria* were used to define three confidence quantiles as used above. Similar levels of goodness of fit were obtained between groups, both regarding first-order (log-likelihood patients: −0.41 ± 0.13; log-likelihood controls: −0.59 ± 0.13) and second-order outcomes (log-likelihood patients: −29.50 ± 1.72; log-likelihood controls: −25.80 ± 1.14). No differences across groups were found for the five parameters (non-decision time: t(35.7) = 0.46, p = 0.75, BF = 0.39; decision bound: t(37.5) = 0.05, p = 0.96, BF = 0.31; drift rate: t(35.1) = 0.01, p = 0.99, BF = 0.31; confidence criterion 1: t(37.6) = −0.04, p = 0.97, BF = 0.31; confidence criterion 2: t(36.2) = −0.29, p = 0.77, BF = 0.32, see Figure 4). In addition, metacognitive efficiency resulting from these simulated data was equivalent in individuals with schizophrenia (mean estimate = 0.52, highest posterior density interval = [0.40 0.65]) and healthy controls (mean estimate = 0.58, highest posterior density interval = [0.40 0.88]), mimicking our behavioral results.

**Figure 1:**
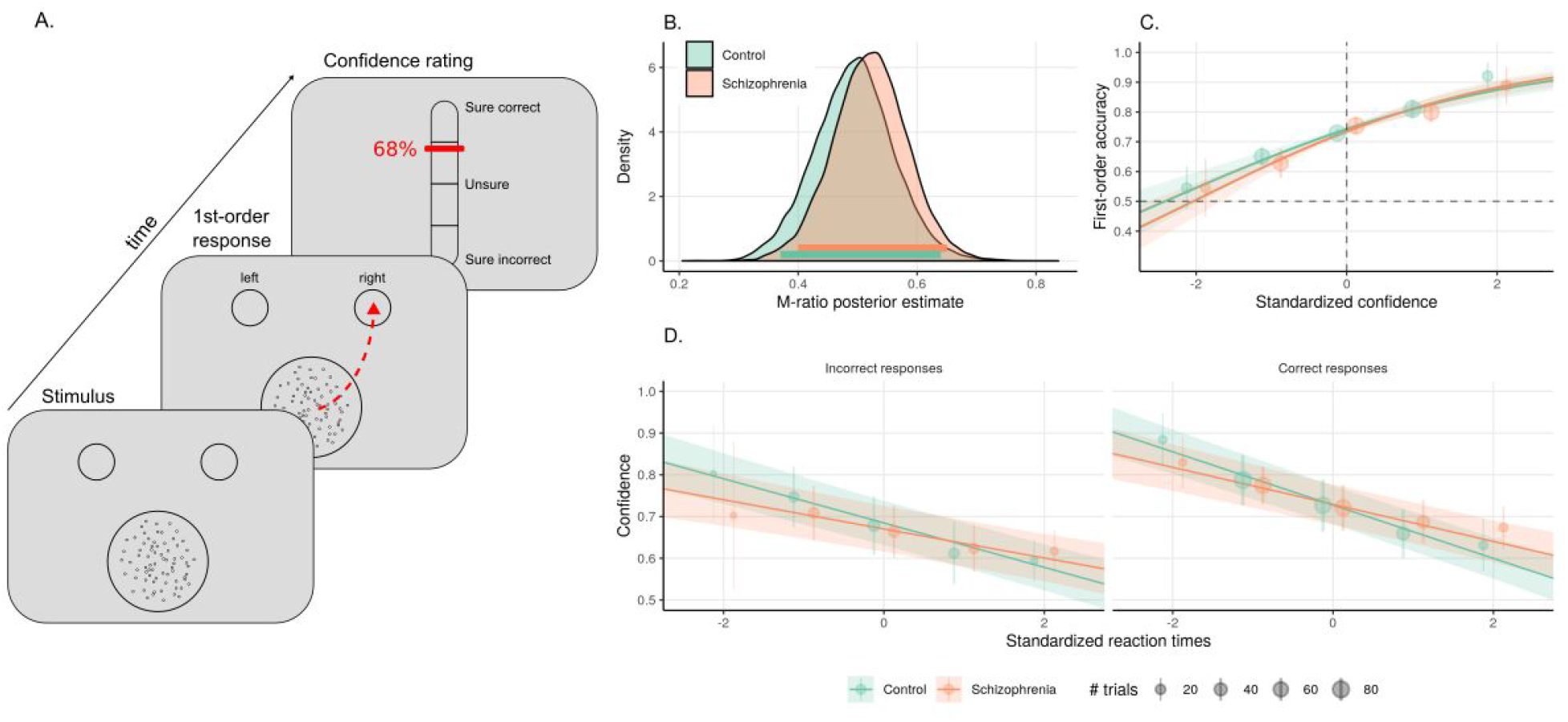
Experimental paradigm and behavioral performance. A. Experimental paradigm. Participants were presented with a random dot kinetogram stimulus moving rightward or leftward and were asked to report motion direction by moving the mouse cursor towards a circle presented at the top-left or top-right of the screen (first-order response). Subsequently, participants reported the confidence they had in their response by moving a cursor on a visual analog scale (second-order response). Exemplar mouse trajectory and confidence ratings are shown in red. B. posterior distribution density of M-Ratio for the control (green) and schizophrenia groups (orange). The colored lines at the bottom of the plot represent the 95% highest posterior density intervals. C. Mixed-effects logistic regression between first-order accuracy and standardized confidence. D. Mixed-effects linear regression between standardized reaction times and confidence. In panels C-D, regression lines and 95 % confidence intervals around them represent the model fit. Although the model took continuous variables as input, we plot for illustrative purposes dots and error bars that represent mean ± 95% confidence interval over participants after rounding standardized confidence (C) and reaction times (D). The size of each dot is proportional to the number of represented trials.

**Figure 3:**
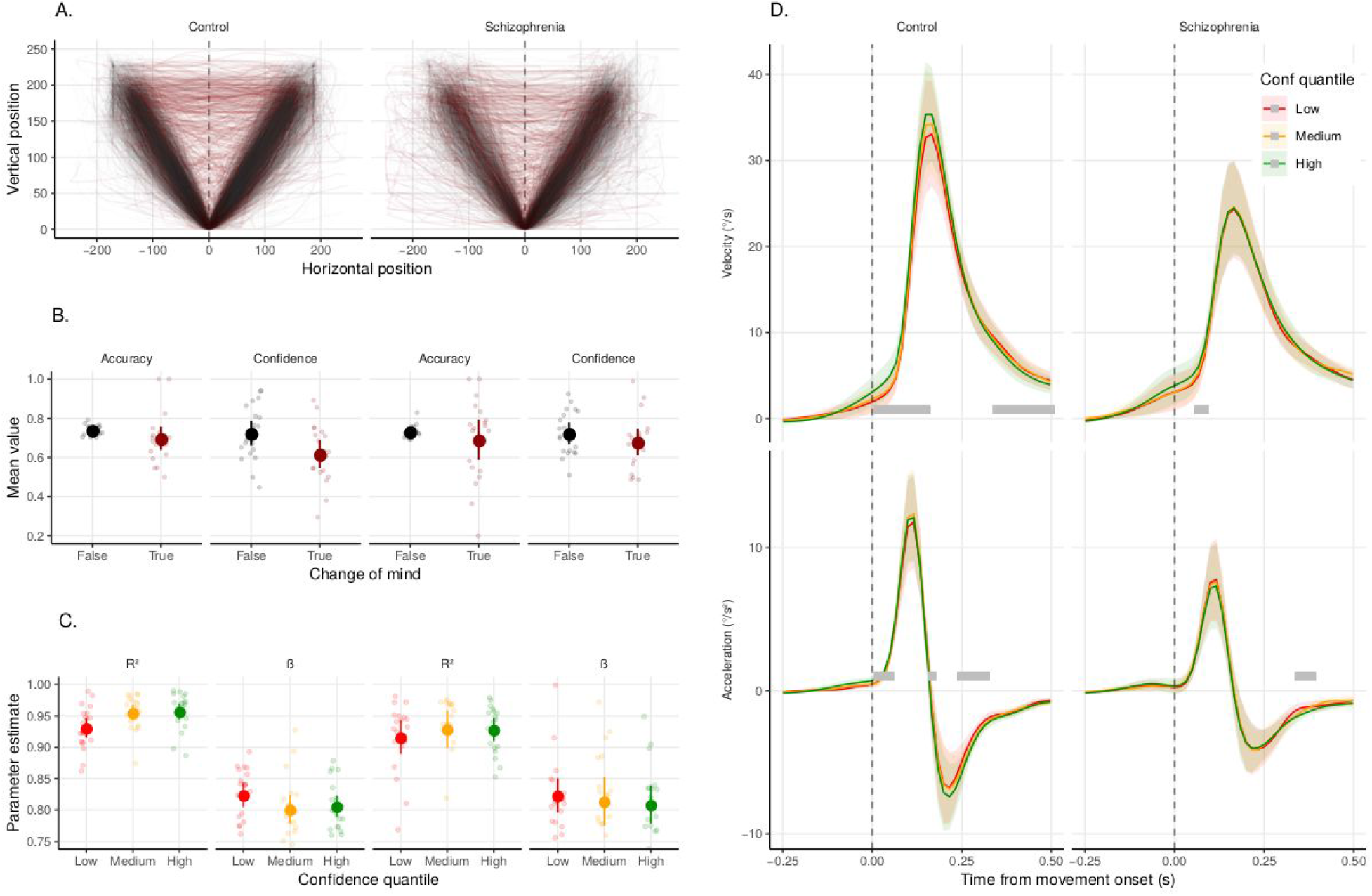
Trajectory tracking. A. Single-trial mouse trajectories leading to the first-order response in case of a change of mind (red) or no change of mind (black) in the control (left panel) and patient groups (right panel). B. Average first-order accuracy and confidence in the presence (red) and absence (black) of a change of mind in the control (left panels) and patient groups (right panels). C. Goodness of fit (R^2^) and slope (β) of the linear fit between vertical and horizontal mouse positions as a function of confidence quantile (low: red, medium: orange, high: green). Large dots represent average estimates, error bars represent the 95 % confidence intervals. Small dots represent individual estimates. D. Average velocity (upper panel) and acceleration (lower panel) from first mouse movement onset as a function of confidence quantile (low: red, medium: orange, high: green). Shaded areas represent the 95 % confidence intervals. Gray bars represent samples for which confidence covaried significantly with kinematics (p < 0.05, fdr-corrected).

**Figure 4:**
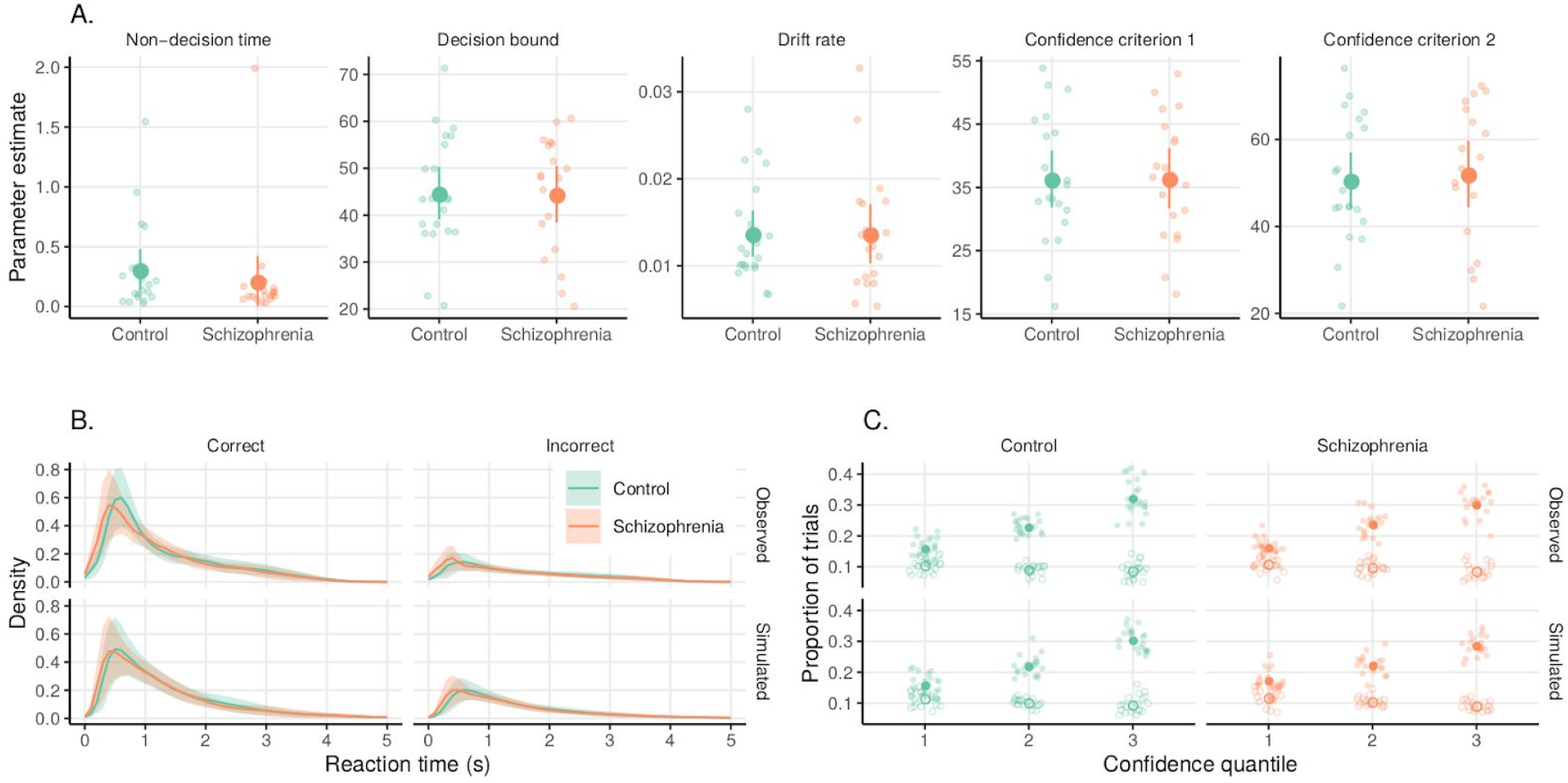
Bounded evidence accumulation model of first and second-order behavior in the control (green) and schizophrenia group (orange). A. Parameter estimates. B. Distributions of observed simulated first-order reaction times for correct and error responses. C. Proportion of trials across confidence quantiles for correct (full dots) and incorrect responses (empty dots). Large dots represent average estimates, error bars represent the 95 % confidence intervals. Small dots represent individual estimates. Shaded areas represent the 95 % confidence intervals.

### 3. Cognitive and clinical variables

The two groups did not differ in terms of gender (χ^2^ = 0.45, p = 0.50), age, education, premorbid intelligence levels, and neuropsychological performance, except for the total score in the Six Elements Test which was marginally lower for patients (mean 801.0 ± 101.4) than in controls (mean 920.8 ± 86.4, t(37.1) = 1.76, p = 0.086, see Table 1). Two other variables differed between patients and controls: depressive symptoms which were higher in patients (mean 0.5 ± 0.4) compared with controls (mean 4.5 ± 1.8, t(21.2) = −4.3, p < 0.001), and cognitive insight scores which were higher in patients (mean 5.8 ± 3.2) than in controls (mean −0.6 ± 1.7, t(28.7) = −3.4, p = 0.002). Of note, the latter difference was not significant anymore when taking into account depression as a covariate: a linear model of insight as a function of group and depression scores revealed a main effect of depression (beta = 0.75 ± 0.30, t(37) = 2.52, p = 0.02, BF = 102.86), but no effect of group (t(37) = 1.55, p = 0.13, BF = 1.21), suggesting that the difference in insight between groups was explained by depression. Depressions scores were >= 6 for 45% of patients indicating a possible major depressive disorder in these participants (Addington, Addington, Maticka-Tyndale, & Joyce, 1992). The CDS total score was < 6 for all participants in the control group. Behavioral results (confidence, B-ROC) remained unchanged when CDS total score was entered as a covariate. In the patients’ sample, the intensity of schizophrenia was measured with the Positive And Negative Syndrome Scale (PANSS). The mean PANSS total score was 78.5 ± 6.8, the mean positive symptoms score was 17.2 ± 2.2, the mean negative symptoms score was 20.5 ± 2.3 and the mean general psychopathology score was 40.9 ± 3.8. The mean illness duration was 14.7 years ± 3.7 and the mean chlorpromazine equivalent was 439.7 mg/24h ± 118.4. The mean score on PSP was 55.7 ± 5.4 and the mean total score on BIS was 10.8 ± 1.1. The group of patients included 13 participants with schizophrenia and 7 with schizo-affective disorders.

**Table 1.** Clinical and neuropsychological characteristics of patients and controls. See methods for details on the psychometric evaluation.

| | Control (N = 21)<br>(mean ± 95%CI) | | Schizophrenia (N = 20)<br>(mean ± 95%CI) | | t-statistic | Degrees of freedom | p-value | Bayes factor |
| --- | --- | --- | --- | --- | --- | --- | --- | --- |
| Age (years) | 42.6 | 4.8 | 38.8 | 5.1 | 1.09 | 37.87 | 0.285 | 0.50 |
| Beck Cognitive Insight Scale | -0.6 | 1.7 | 5.8 | 3.2 | -3.39 | 28.74 | 0.002 | 20.96 |
| Calgary Depression Scale | 0.5 | 0.4 | 4.5 | 1.8 | -4.26 | 21.21 | 0.001 | 171.20 |
| Education level (years) | 12.5 | 0.4 | 13.6 | 1.3 | -1.56 | 22.70 | 0.133 | 0.80 |
| Premorbid IQ | 104.0 | 3.6 | 102.3 | 3.9 | 0.65 | 37.74 | 0.521 | 0.37 |
| Six Elements Test (errors) | 9.2 | 1.4 | 8.0 | 2.1 | 0.93 | 32.94 | 0.359 | 0.44 |
| Six Elements Test (points) | 920.8 | 86.4 | 801.0 | 101.3 | 1.76 | 37.07 | 0.086 | 1.04 |
| WAIS matrix subtest | 10.2 | 1.1 | 9.0 | 1.3 | 1.33 | 37.13 | 0.192 | 0.62 |
| WAIS letter-number sequencing subtest | 9.1 | 1.2 | 7.7 | 1.1 | 1.69 | 37.98 | 0.100 | 0.94 |
| WAIS vocabulary subtest | 10.0 | 1.2 | 11.1 | 1.5 | -1.06 | 36.54 | 0.297 | 0.48 |

We examined the relationship between second-order behavioral measures (confidence, M-Ratio) and these cognitive and clinical variables using Bayesian robust regressions (see methods). Regarding cognitive variables, we found that M-Ratio covaried positively with the WAIS matrix subtest (r = 0.46, HDP = [0.20 0.70], Bayes Factor = 13.88) on the whole group of participants, indicating that participants with good perceptual reasoning also had high metacognitive performance. No other correlation was found significant (see Supplementary Table S1). No significant correlation was found between second-order behavioral measures and clinical variables specific to the patient population (see Supplementary Table S2).

## Discussion

The current study sought to systematically assess the quality of metacognitive monitoring in perceptual decision-making in schizophrenia spectrum disorders, using bias-free measures of metacognitive performance combined with trajectory tracking and bounded evidence accumulation models of decision-making.

### Metacognitive performance

No significant difference in metacognitive efficiency and sensitivity was found between groups, with Bayesian analyses favoring the null hypothesis rather than inconclusive results. In addition, a bounded evidence accumulation model suggested that both groups relied on equivalent first and second-order decisional mechanisms when compensating for first-order perceptual deficits in patients. The absence of a difference between groups is unlikely due to abnormally poor metacognitive performance in our control population, as healthy participants performed similarly to participants from previous studies involving a coherent motion discrimination paradigm (e.g., mean M-Ratio = 0.66 in Konishi, Compain, Berberian, Sackur, & de Gardelle, 2019). Plus, the unexpected worse cognitive insight found in patients compared to controls can not explain the lack of difference in metacognitive performance between groups since cognitive insight was related neither with metacognitive efficiency nor metacognitive sensitivity. One might argue that including individuals with schizo-affective disorders might have compensated for a potential deficit in metacognition in individuals with schizophrenia, by increasing depressive symptomatology, which has been associated with better metacognitive efficiency (Rouault, Seow, Gillan, & Fleming, 2018). However, the prevalence of possible depression (based on the CDS cut-off) in this sample of individuals with schizophrenia spectrum disorders was close to the 40% value reported in another study investigating outpatients with schizophrenia only (an der Heiden, Leber, & Häfner, 2016).

Recent studies reported lower visual metacognitive performance in individuals with a first episode of psychosis relative to age-matched controls (Alkan, Davies, Greenwood, & Evans, 2019; Davies et al., 2018). Here, we tested individuals with a chronic disorder and found no deterioration of metacognition relative to controls, but a negative relationship between metacognitive efficiency and illness duration (Table S2). This suggests that metacognitive performance in patients may evolve non-linearly over time, with prevalent deficits at the early and late stages of schizophrenia. Longitudinal studies with large sample sizes and varying ages of onset will be needed to assess this possibility.

Regarding chronic schizophrenia, a study reported lower metacognitive performance in a source memory task compared to psychiatric control groups (obsessive-compulsive and post-traumatic stress disorders) with similar type-1 accuracy (Moritz & Woodward, 2006). However, our results are in line with previous studies investigating metacognitive performance controlling for first-order accuracy in chronic schizophrenia, which reported equivalent metacognitive sensitivity (area under the type 2 ROC curve) between patients and controls for facial emotion recognition (Pinkham, Klein, Hardaway, Kemp, & Harvey, 2018), comparable metacognitive sensitivity (strength of the association between first-order accuracy and confidence) for episodic memory (Bacon & Izaute, 2009), and comparable metacognitive efficiency (M-Ratio) during a detection task (Powers, Mathys, & Corlett, 2017). In contrast, many studies reported a metacognitive deficit in chronic schizophrenia without controlling for concomitant lower first-order performance (Eifler et al., 2015; Köther et al., 2012; Moritz et al., 2008; Moritz et al., 2012). Therefore, an important aspect of future studies quantifying confidence and metacognitive performance in schizophrenia will be to systematically control for potential confounds in terms of first-order performance.

### Relationships between action execution and metacognitive performance

Reaction times were not longer in individuals with schizophrenia spectrum disorders, nor were mouse movement onsets, maybe as a consequence of the artefactual matching of task difficulty and accuracy between the two groups. Both groups featured a negative relationship between reaction times and confidence, but significantly stronger in the control group. This result is in line with a previous report showing a lack of correlation between reaction times and confidence in emotion recognition for individuals with schizophrenia whereas reaction times were negatively associated with confidence in controls (Jones et al., 2019). Together, these results suggest that decisional parameters such as reaction times have less influence on subsequent confidence ratings in individuals with schizophrenia spectrum disorders.

The analysis of reaction times with a bounded evidence accumulation model revealed no difference in parameters between groups. This is at odds with the first study employing this technique in schizophrenia, showing that patients had an increased non-decisional time, higher boundary separation and lower drift rate in a punishment learning task (Moustafa et al., 2015). Longer non-decisional time and lower drift rate were also recently found for speeded judgments among individuals with schizophrenia and their siblings compared to healthy controls (Fish et al., 2018).

Beyond mere reaction times, we found that velocity and acceleration during the decision movement were more closely linked to confidence in the control than in the schizophrenia group. The link between confidence and trajectories in the control group corroborates the view that sensorimotor signals shape confidence estimates. Indeed, previous studies showed that electromyographic activity (Gajdos, Fleming, Saez Garcia, Weindel, & Davranche, 2019) and alpha power over somatosensory scalp regions (Faivre, Filevich, Solovey, Kuhn, & Blanke, 2018) covary with confidence, and that altering sensorimotor signaling by increasing movement speed (Palser, Fotopoulou, & Kilner, 2018) or by inducing sensorimotor conflicts (Faivre, Vuillaume, et al., 2019) disrupt metacognitive accuracy. The weaker link between trajectories and confidence in schizophrenia may be related to slower and noisier motor behavior, or to the tendency of patients to neglect relevant internal cues to control motor actions (Frith, Blakemore, & Wolpert, 2000). The fact that metacognitive performance was preserved in schizophrenia despite a decreased link between confidence and trajectories suggest that sensorimotor signals may globally up or down-regulate confidence estimates, with no influence on the calibration between confidence and first-order performance as reported recently (Filevich, Koß, & Faivre, 2019).

### Relationships between behavioral and neuropsychological outcomes

No difference was found between patients and controls according to premorbid IQ, perceptual and verbal reasoning and working memory. Executive functions were marginally lower in patients. In contrast, coherent motion discrimination was significantly worse in patients compared to controls in line with previous studies (Chen, 2011) thus suggesting a deficient integration of spatially distributed motion signals in patients. In the group of patients, schizophrenic symptomatology was moderate (Leucht et al., 2005) and the level of depression slightly higher than what is usually reported in stabilized outpatients sample (Roux et al., 2018; Tanaka et al., 2012). Depressive symptomatology was also higher in patients than in controls for the current study. Patients reported mean clinical (Ehrminger et al., 2019) and cognitive (Kim, Lee, Han, Kim, & Lee, 2015; Misdrahi, Denard, Swendsen, Jaussent, & Courtet, 2014; Phalen, Viswanadhan, Lysaker, & Warman, 2015) insights which were comparable to those reported in previous studies including stabilized outpatients. In contrast, cognitive insight was markedly lower in the controls we had recruited compared to previous studies (Kao & Liu, 2010; Martin, Warman, & Lysaker, 2010; Uchida et al., 2009), which however included much younger and educated non-clinical participants than the ones included in the current sample. The higher level of depression found in patients compared to controls and the low cognitive insight found in controls regarding the level usually reported both converged to explain that cognitive insight was unexpectedly better in patients than in controls. This difference was indeed not significant anymore when depression was entered as a covariate. We found a significant correlation between metacognitive efficiency and visual reasoning on the whole group of participants, which suggests that metacognition and reasoning abilities depend on partially overlapping cognitive mechanisms (Mäntylä, Rönnlund, & Kliegel, 2010). Previous studies reported that metamemory correlated with executive function, visual recognition memory (Chiu, Liu, Hwang, Hwu, & Hua, 2015) and working memory (Eifler et al., 2015). Contrary to a previous study reporting a significant association between poor insight and metacognitive deficits in schizophrenia (Koren et al., 2004), we found no correlation between illness insight and metacognitive performance or confidence bias. Plus, metacognitive performance did not correlate with psychosocial functioning in patients. Our study thus does not confirm the significant association between synthetic metacognition (drawing upon a broad range of social, executive, linguistic, and metacognitive processes, such as the Metacognitive Assessment Scale) and functioning previously reported (Arnon-Ribenfeld et al., 2017).

### Conclusions

Controlling for the accuracy of first-order decisions and other important cognitive confounds like perceptual reasoning, we found equivalent decision-making and metacognitive processes in individuals with schizophrenia and healthy matched controls. However, confidence was less related to the speed of decision and to motor response parameters like velocity and acceleration in individuals with schizophrenia. This study emphasizes the importance to run future studies controlling for first-order accuracy and reasoning before concluding that individuals with schizophrenia have a specific metacognitive deficit.

## Methods

The experimental paradigm and analysis plan detailed below were registered prior to data collection (NCT03140475) and are available together with anonymized data and analyses scripts on the open science framework (https://osf.io/84wqp/).

### Participants

Individuals with schizophrenia spectrum disorders (schizophrenia or schizoaffective disorder) were recruited from community mental health centers and outpatient clinics in the Versailles area. The control participants were recruited from the volunteers’ panel at the *Centre d’Economie de la Sorbonne* and Versailles Hospital. Exclusion criteria for both groups were a moderate-to-severe substance use disorder (DSM-5 criteria) within the 12 months preceding the study, and a current or prior untreated medical illness, including neurologic illness, an IQ < 70 based on three subtests of the Wechsler Adult Intelligence Scale (see below), and an age > 60 years. Schizophrenia and schizoaffective disorders were diagnosed by M.R. based on the Structured Clinical Interview for assessing the DSM-5 criteria (First, Williams, Karg, & Spitzer, 2016). Another licensed psychiatrist (patient’s treating psychiatrist) confirmed the diagnosis for each patient according to the DSM-5 criteria. The control group was screened for current or past psychiatric illness, and individuals were excluded if they met the criteria for a severe and persistent mental disorder.

All participants were right-handed, had normal hearing and normal or corrected-to-normal vision. They were naive to the purpose of the study and gave informed consent. The study was approved by the ethical committee *Sud Méditérannée* II (217 R01). Our plan at pre-registration was to collect data until we reach a Bayes Factor of either 1/3 or 3. We halted data collection when evidence for the null hypothesis in our main test of interest was obtained (i.e., M-Ratio, see analyses below).

### Neuropsychological and clinical evaluation

Both individuals with schizophrenia spectrum disorders and healthy controls were evaluated on the following neuropsychological domains :

- perceptual reasoning with the standardized score on the matrices subtest of the Wechsler Adult Intelligence Scale 4th version (WAIS-IV, Wechsler, Coalson, & Raiford, 2008)
- verbal reasoning with the standardized score on the vocabulary subtest of WAIS-IV
- working memory with the standardized score on the letter-number sequencing subtest of WAIS-IV
- executive functions with the raw total and error scores on the Modified Six Elements Test (Wilson, Evans, Alderman, Burgess, & Emslie, 1997)
- depressive symptoms with the Calgary Depression Scale (CDS) (Addington et al., 1992)
- cognitive insight with the composite index on the Beck Cognitive Insight Scale (BCIS) (Beck, Baruch, Balter, Steer, & Warman, 2004). The composite index of the BCIS reflects the cognitive insight and is calculated by subtracting the score for the self-certainty scale from that of the self-reflectiveness scale.
- the National Adult Reading Test (NART) provided an estimate of premorbid IQ (Nelson & O’Connell, 1978)

The following clinical evaluations were run for patients only:

- the intensity of schizophrenia symptoms with the Positive And Negative Syndrome Scale (Kay, Fiszbein, & Opfer, 1987)
- social functioning using the Personal and Social Performance Scale (PSP) (Morosini, Magliano, Brambilla, Ugolini, & Pioli, 2000)
- clinical insight using the Birchwood insight scale (Birchwood et al., 1994)

### Procedure

Among multiple perceptual paradigms, coherent motion discrimination was chosen because it had been recommended as a promising perception paradigm for translation for use in clinical trials due to good psychometric validity (Green et al., 2008). All stimuli were prepared and presented using the psychophysics toolbox under Matlab (Brainard, 1997; Kleiner et al., 2007; Pelli, 1997), based on a previous study (de Gardelle & Mamassian, 2015). Participants started each trial by clicking on a 1.2° × 3.6° box placed at the bottom of the screen (Figure 1). The mouse click triggered the display of a 0.1° fixation dot presented in the middle of a circular frame (3° radius) for 250 ms over a gray background, followed by the display of a random dot kinetogram (RDK), consisting of 100 dots (radius 0.1°) moving pseudo randomly at a constant speed of 3°/s. The motion direction of each dot was drawn every 16 ms from a von Mises distribution that determined both the mean (± 45° relative to the vertical) and variance of the motion direction. Visual transients at stimulus onset were smoothed using a linear ramp in contrast. On each trial, participants were asked to indicate the mean motion direction by clicking within a circular frame (radius 1.35°) located on the top, to the right or to the left of the RDK. Responses slower than 6 s were discouraged by playing a loud alarm sound. The task difficulty was adjusted by a one-up two-down staircase procedure to make the first-order performance rate converge to 71 % (Levitt, 1971). Perceptual evidence was defined as the inverse of variance in motion direction, increasing after one incorrect response and decreasing after two consecutive correct responses. After providing their first-order response, a visual analog scale appeared, and participants were asked to report how confident they were about it by moving a slider vertically using the mouse. The scale was presented until a response was provided, with marks between 0 % (certainty that the first-order response was erroneous) and 100 % (certainty that the first-order response was correct) with 5% steps. The initial position of the cursor was always 50 %. The experiment was divided into 10 blocks of 30 trials and lasted about 1 hour. The perceptual difficulty was pre-tuned to individual perceptual abilities by performing 80 trials without confidence ratings prior to the main experiment.

### Statistical analysis

All analyses were performed with R (2018). We compared groups’ characteristics using the Welch t-test or χ2 test when appropriate. Trials in which reaction times were above 6 s or below 200 ms were excluded (first-order responses: 2.2 ± 0.9 % of total trials; second-order response 1.5 ± 0.7 %). All Bayesian models were created in Stan computational framework (http://mc-stan.org/) accessed with the brms package (Bürkner, 2017), based on four chains of 10000 iterations including 2000 warmup samples. We report the highest density probability for all estimates, which specifies the range covering the 95% most credible values of the posterior estimates. Mixed-effects models included random intercepts by participants and full random effects structure. Metacognitive sensitivity was quantified using a mixed-effects logistic regression between first-order accuracy and confidence, including a fixed effect of group (controls vs. patients). Metacognitive efficiency was quantified in a Bayesian framework as the ratio between meta-d’ and d’ (M-Ratio) (Fleming, 2017; Maniscalco & Lau, 2012). Confidence bias quantified differences in the tendency to use high or low confidence ratings. It was calculated based on the second-order receiver operating characteristic curve (ROC) which determines the rate of correct and incorrect responses at given confidence levels. Namely, confidence bias corresponded to the log-ratio between the lower and upper area separated by the minor diagonal (B-ROC) (Kornbrot, 2006). Mouse spatial trajectories (X, Y), and kinematics (instantaneous velocity and acceleration) associated with first-order responses were smoothed using a Savitzky-Golay filter of order 2 and length 5. Trajectories were temporally realigned with respect to movement onset, defined as the time when velocity reached 20% of the maximal velocity in a given trial. Trajectories were spatially realigned with respect to the spatial coordinate at movement onset (i.e., starting point). Spatial coordinates were fitted using a model II linear regression with the major axis method (Legendre, 2018). Kinematics (velocity, acceleration) were standardized across participants (z-score) and analyzed as a function of confidence using mixed-effects linear regressions. Changes of mind were defined as trials in which the sign of the trajectory angle at movement onset was opposite to that of the landing point (i.e, when participants started moving towards one side of the screen but ended responding on the other side). Only initial angles with an absolute value between 10 and 80 degrees were considered to exclude trials with vertical or horizontal trajectories at movement onset. Robust Bayesian correlations were computed using Stan with 10000 iterations including 2000 warmup samples and non-informative priors, assuming that pairs of psychometric and behavioral outcomes followed a bivariate Student’s t-distribution.

### Bounded accumulation model

To reduce the degrees of freedom of the model, we fixed the standard deviation of the non-decision time to 60 ms, the standard deviation of the within-trial noise to 1 and the correlation of the two accumulators to -sqrt(0.5) as in previous works (e.g. Van Den Berg et al., 2016). The accumulation process was bounded negatively to zero. We used Euler’s method to simulate 1000 trials of the evidence accumulation process and the corresponding response times (i.e. movement onsets) and choice accuracies (i.e. accuracy of the movement direction at the onset), which were fitted to the observed data for every participant. For this, we maximized the log-likelihood using a Nelder-Mead simplex method. The log-likelihood was computed using a two-sample Kolmogorov-Smirnov test on the simulated and observed movement onset times, inverting their sign when the direction was incorrect. We repeated this procedure with a wide range of initial parameters to avoid local minima. We fitted confidence ratings in a second stage; we simulated 1000 paths of the evidence accumulation process and discretized the winning accumulator (at a fixed readout time after the decision) into three confidence levels using two criteria. The values of the two criteria were initialized to approximate the proportion of high and low confidence ratings and then fitted to the data using a Nelder-Mead simplex method. For this, we computed the log-likelihood using a Bernoulli probability distribution for each confidence level and choice accuracy. We repeated this procedure using readout timings going from 0 s after the initial decision to 1 s after the initial decision by steps of 100 ms and used the model with the best likelihood. Of note, our results still held when using fixed readout times across participants or when fitting confidence ratings to the state of the loosing accumulator (Kiani & Shadlen, 2009) or to the difference between the winning and losing accumulator (*balance-of-evidence* (Rahnev, Nee, Riddle, Larson, & D’Esposito, 2016; Vickers, 1979).

## Data Availability

The experimental paradigm and analysis plan detailed below were registered prior to data
collection ( NCT03140475 ) and are available together with anonymized data and analyses
scripts on the open science framework ( https://osf.io/84wqp/ ).

## Acknowledgments

We thank Eric Brunet for his pertinent comments about the interpretation of our results. We are also grateful to Laure Morisset at Versailles Hospital for her institutional support.

N.F. received funding from the European Research Council under the European Union’s Seventh Framework Programme (FP/2007-2013) / ERC Grant Agreement n. 803122.

## Supplementary materials

**Figure S1.**
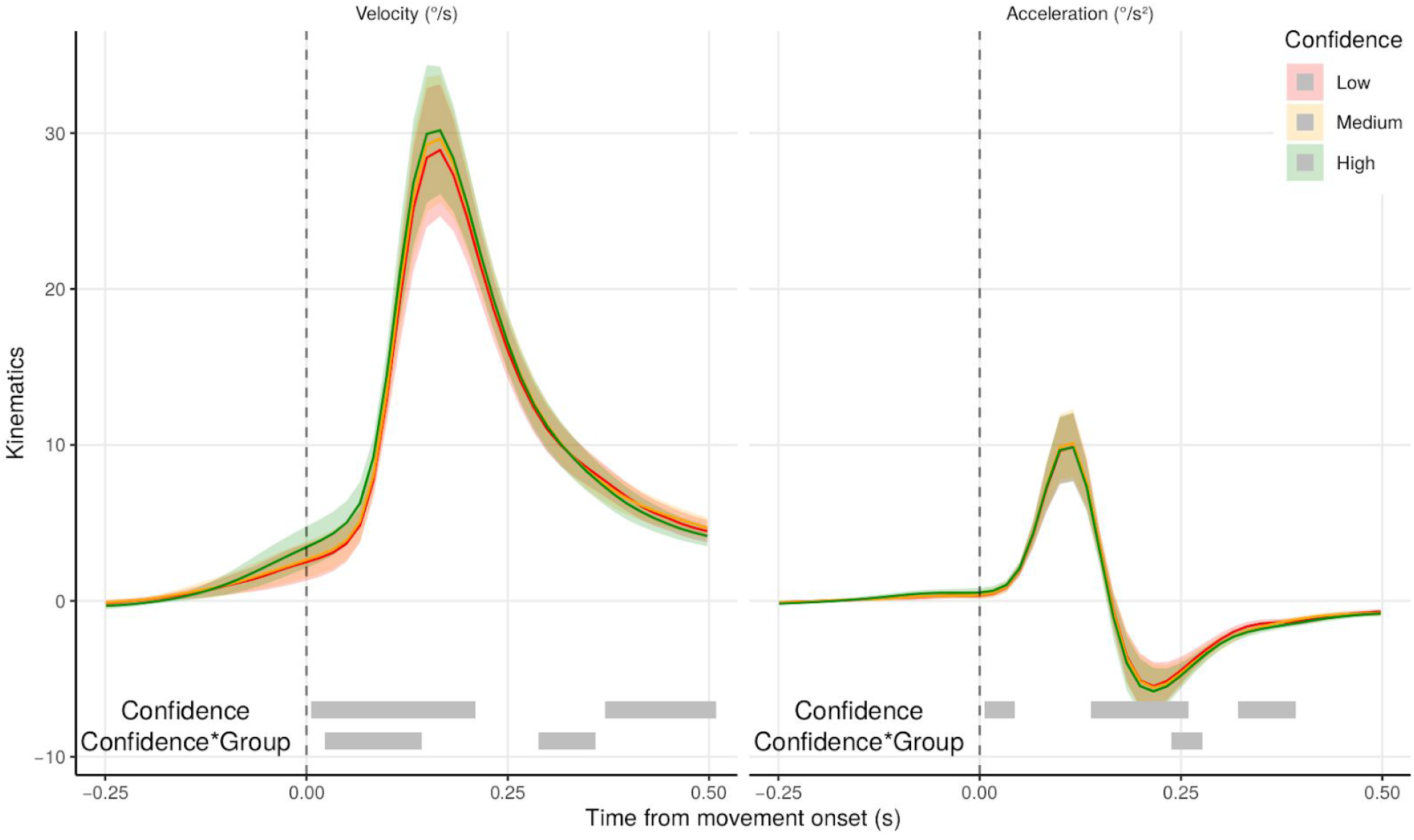
Average velocity (left panel) and acceleration (right panel) from first mouse movement onset as a function of confidence quantile (low: red, medium: orange, high: green). Shaded areas represent the 95 % confidence intervals. Gray bars represent samples with main effects of confidence or interaction between confidence and group kinematics (p < 0.05, fdr-corrected).

**Table S1:** Correlations between behavioral results and neuropsychological characteristics of individuals with schizophrenia spectrum disorders and controls.

| Covariates |  | Correlation coefficient | HPD |  | Posterior probability | Bayes Factor |
| --- | --- | --- | --- | --- | --- | --- |
| Confidence | Age (years) | 0.19 | -0.14 | 0.48 | 0.123 | 0.33 |
|  | Beck Cognitive Insight Scale | -0.29 | -0.58 | 0.04 | 0.046 | 0.75 |
|  | Calgary Depression Scale | -0.19 | -0.48 | 0.14 | 0.137 | 0.33 |
|  | Education level (years) | -0.08 | -0.40 | 0.25 | 0.315 | 0.20 |
|  | Premorbid IQ | 0.19 | -0.12 | 0.50 | 0.129 | 0.31 |
|  | Six Elements Test (errors) | 0.29 | -0.01 | 0.57 | 0.036 | 0.83 |
|  | Six Elements Test (points) | -0.06 | -0.39 | 0.26 | 0.356 | 0.19 |
|  | WAIS matrix subtest | -0.37 | -0.63 | -0.07 | 0.014 | 2.42 |
|  | WAIS mequence subtest | 0.04 | -0.30 | 0.36 | 0.402 | 0.18 |
|  | WAIS vocabulary subtest | 0.13 | -0.20 | 0.44 | 0.228 | 0.23 |
| M-Ratio | Age (years) | -0.25 | -0.54 | 0.06 | 0.064 | 0.54 |
|  | Beck Cognitive Insight Scale | 0.17 | -0.15 | 0.48 | 0.159 | 0.29 |
|  | Calgary Depression Scale | 0.18 | -0.16 | 0.47 | 0.140 | 0.31 |
|  | Education level (years) | 0.20 | -0.12 | 0.50 | 0.122 | 0.34 |
|  | Premorbid IQ | 0.25 | -0.05 | 0.54 | 0.061 | 0.55 |
|  | Six Elements Test (errors) | -0.22 | -0.53 | 0.08 | 0.088 | 0.46 |
|  | Six Elements Test (points) | 0.28 | -0.03 | 0.55 | 0.046 | 0.71 |
|  | WAIS matrix subtest | 0.46 | 0.20 | 0.70 | 0.002 | 13.88 |
|  | WAIS mequence subtest | 0.33 | 0.02 | 0.59 | 0.022 | 1.45 |
|  | WAIS vocabulary subtest | 0.32 | 0.02 | 0.58 | 0.024 | 1.13 |

**Table S2:** Correlations between behavioral results and clinical characteristics of individuals with schizophrenia spectrum disorders.

| Covariates |  | Correlation coefficient | HPD |  | Posterior probability | Bayes Factor |
| --- | --- | --- | --- | --- | --- | --- |
| Confidence | Birchwood Insight Scale | 0.12 | -0.33 | 0.57 | 0.315 | 0.29 |
|  | Chlorpromazine equivalent | 0.17 | -0.30 | 0.57 | 0.233 | 0.32 |
|  | Illness duration | 0.27 | -0.17 | 0.66 | 0.118 | 0.51 |
|  | PANSS Negative symptoms score | 0.05 | -0.38 | 0.52 | 0.416 | 0.25 |
|  | PANSS Positive symptoms score | 0.36 | -0.06 | 0.73 | 0.065 | 0.84 |
|  | PANSS General psychopathology score | -0.22 | -0.62 | 0.25 | 0.183 | 0.37 |
|  | PANSS total score | 0.01 | -0.44 | 0.46 | 0.486 | 0.25 |
|  | Personal and Social Performance Scale | -0.22 | -0.63 | 0.22 | 0.175 | 0.39 |
| M-Ratio | Birchwood Insight Scale | 0.13 | -0.34 | 0.54 | 0.299 | 0.29 |
|  | Chlorpromazine equivalent | -0.15 | -0.56 | 0.32 | 0.266 | 0.30 |
|  | Illness duration | -0.54 | -0.83 | -0.18 | 0.006 | 5.22 |
|  | PANSS Negative symptoms score | -0.31 | -0.67 | 0.15 | 0.095 | 0.55 |
|  | PANSS Positive symptoms score | -0.19 | -0.60 | 0.28 | 0.223 | 0.34 |
|  | PANSS General psychopathology score | 0.03 | -0.43 | 0.47 | 0.450 | 0.24 |
|  | PANSS total score | -0.13 | -0.54 | 0.32 | 0.295 | 0.28 |
|  | Personal and Social Performance Scale | 0.31 | -0.13 | 0.69 | 0.090 | 0.61 |

## Notes

### Competing Interest Statement

The authors have declared no competing interest.

### Clinical Trial

NCT03140475

